# Comparative complete scheme and booster effectiveness of COVID-19 vaccines in preventing SARS-CoV-2 infections with SARS-CoV-2 Omicron (BA.1) and Delta (B.1.617.2) variants

**DOI:** 10.1101/2022.01.31.22270200

**Authors:** Irina Kislaya, André Peralta-Santos, Vítor Borges, Luís Vieira, Carlos Sousa, Bibiana Ferreira, Ana Pelerito, João Paulo Gomes, Pedro Pinto Leite, Baltazar Nunes, PT COVID-19 group

## Abstract

**Introduction:** Information on vaccine effectiveness and viral loads in a context of novel variants of concern (VOC) emergence is of key importance to inform public health policies. This study aimed to estimate a measure of comparative vaccine effectiveness between Omicron (BA.1) and Delta (B.1.617.2 and sub-lineages) VOC according to vaccination exposure (primary or booster) and time since primary vaccination and to compare cycle threshold (Ct) values between Omicron and Delta VOC infections according to the vaccination status as an indirect measure of viral load.

**Methods:** We developed a case-case study using data on RT-PCR SARS-CoV-2 positive cases notified in Portugal during weeks 49-51 2021. The odds of vaccination in Omicron cases were compared to Delta using logistic regression adjusted for age group, sex, region and week of diagnosis and laboratory of origin. RT-PCR Ct values were compared by vaccination status and variant using linear regression model.

**Results:** Higher odds of vaccination were observed in cases infected by Omicron (BA.1) VOC compared to Delta (B.1.617.2) VOC cases for both complete primary vaccination (OR=2.1; CI 95% :1.8 to 2.4) and booster dose (OR= 5.2; CI 95%: 3.1 to 8.8), indicating vaccine effectiveness reduction against Omicron. No differences in distribution of Ct-values between these two VOC were observed for any vaccination exposure categories.

**Conclusion:** Consistent lower VE was observed against Omicron infection. Complete primary vaccination may not be protective against SARS-CoV-2 infection in regions where Omicron variant is dominant, but a massive rollout of booster vaccination campaign can contribute to reduce SARS-CoV-2 incidence in the population.

## Background

The Omicron (BA.1) SARS-CoV-2 variant first reported in South Africa on 24 November 2021(1) has been classified by the World Health Organization(2) as a variant of concern (VOC), as it presents several mutations associated with increased transmissibility, immune escape, and higher risk of reinfection(3,4). The Omicron (BA.1) variant yields an S gene target failure (SGTF) signal (due to the deletion Δ69–70 in the Spike protein) in some PCR tests (e.g. TaqPath COVID-19, ThermoFisher, Waltham, MA, United States), which can be used as a proxy for Omicron detection and differentiation from the circulating Delta (B.1.617.2 and sub-lineages) variant (in which the Δ69–70 is rarely detected)(5).

The Omicron had a swift rise in Europe becoming dominant in a few weeks in England, Scotland and Denmark(6), and the European Center for Disease Prevention and Control (ECDC) risk assessment(7) referred that would become dominant in early January 2022 in all European Union member states. Considering COVID-19 vaccination, most European countries have ongoing mass population vaccination, with considerably high vaccine coverage in some countries. Hence, it is fundamental to understand how this novel VOC impacts the transmission dynamics in highly vaccinated populations. The first studies on neutralization assays revealed an extensive but incomplete escape of Cominarty BNT162b2 elicited neutralization(8), but booster dose increased the neutralization(9). These preliminary in-vitro results were confirmed by the first vaccine effectiveness studies in England and Scotland(10,11) for symptomatic infections and hospitalizations for both complete vaccination schemes and booster doses. UK study estimated a reduction of vaccine effectiveness (VE) for symptomatic infection with the Omicron with no effect for two ChAdOx1 (Vaxzevria) doses and 34% and 37% from 15 weeks post dose 2 of BNT162b2 (Cominarty)(10). Another study in Denmark found similar results with reduced VE for Omicron infection compared with Delta for two doses of BNT162b2 (Cominarty) and mRNA-1273 (Spikevax), with an increase in VE after a booster dose(12). It is still unclear the reasons for differences in VE of different vaccines and if the decreased VE translate for other countries with different vaccination coverage.

To shed some light on those questions we aim to replicate a study previously performed to estimate comparative vaccine effectiveness of mRNA vaccines between Delta (B.1.617.2 and sub-lineages) and the Alpha (B.1.1.7) VOC(13). Our objective is to measure comparative vaccine effectiveness (any vaccine) between Omicron and Delta VOC cases according to the complete primary vaccine specific scheme, time since primary vaccine scheme and booster dose, and also compare Cycle threshold (Ct) values between Omicron and Delta VOC according to the vaccination status of the cases. Additionally, we intend to translate comparative vaccine effectiveness estimates between Omicron versus Delta VOC into vaccine effectiveness estimates against Omicron variant infection using published estimates of COVID-19 vaccine effectiveness against Delta.

## Methods

### Study design

To estimate a measure of comparative COVID-19 vaccine effectiveness of complete primary vaccination scheme and of the booster dose against the SARS-CoV-2 Omicron (BA.1) versus Delta (B.1.617.2 and sub-lineages) VOC we used a case-case study design(14). Similar approach was used and has been previously described elsewhere(13). This design has been proven useful to address question about COVID-19 vaccine effectiveness in a context of novel VOC emergence comparing directly the odds of vaccination between RT-PCR–positive cases infected with different VOC. Considering cases of the Omicron SARS-CoV-2 infection as cases of interest, and Delta infections as the reference group, higher vaccination odds in Omicron cases in a case-case study are indicative of a lower effectiveness of COVID-19 vaccines against the Omicron comparative to the Delta VOC.

For the second objective, we have used TaqPath Ct values for nucleocapsid (N) and open reading frame 1ab (ORF1ab) gene as a proxy to compare viral loads for Omicron and Delta cases by vaccination status(15). Ct values indicate a number of RT-PCR cycles required for amplification of SARS-CoV-2 RNA and have been used to infer infectiousness and transmissibility on others VOC(16).

The study period covered 3 weeks (6-26 December 2021), starting on week 49 with predominant circulation of the Delta VOC and relative frequency of the Omicron of 4.2% (17) until week 51, when the Omicron became predominant 50.8%(18).

Target population included individuals aged 12 or more years old, resident in Portugal mainland eligible for vaccination(19) with positive RT-PCR notified to the mandatory National Epidemiological Surveillance Information System (SINAVE) during the study period. To evaluate effect of primary vaccination, data was restricted to individuals without history of previous SARS-CoV-2 infection or vaccine booster. To access the effect of the booster dose, we restricted the sample to those aged 50 or more years old since younger age groups were not yet eligible for the booster vaccination at the time of the study and to those without history of previous SARS-CoV-2 infection(19). Individuals with missing data on National Health Service User number, age, sex, place of residence or diagnosis date were excluded from all analyses.

### Data Sources

#### SARS-CoV-2 Cases

We linked laboratory data on Ct values and whole-genome sequencing on SARS-CoV-2 positive cases collected by the National SARS-CoV-2 Genomic Surveillance Network and three private molecular biology laboratories (UNILABS, Algarve Biomedical Center and Portuguese Red Cross) to the national electronic vaccination register (VACINAS) and the National Epidemiological Surveillance Information System that contains basic demographic information on cases and data of previous SARS-COV-2 infections since the beginning of the pandemic. Deterministic data linkage was performed on 4 January 2022 by the General Directorate of Health using National Health Service User number that uniquely identifies individuals in all national administrative health registries. Based on National Health Service User number, records for duplicated samples were removed maintaining only data from the first collected sample registry for each SARS-CoV-2 infection.

### Variant classification

SARS-CoV-2 variants were classified by viral whole-genome sequencing (WGS) or inferred for non-sequenced samples based on data on S-gene amplification using the TaqPath™ Covid 19 CE IVD RT-PCR Kit (Thermo Scientific™) assay, as follows: no S-gene amplification (Omicron BA.1) and S-gene amplification (Delta). TaqPath S-positive samples could be confidently classified as Delta, since this VOC was dominant in Portugal since the last Summer (weekly frequencies above 99% between weeks 30 and 47, when Omicron started emerging)(https://insaflu.insa.pt/covid19/).

### Vaccination status

In Portugal four vaccines are authorized for primary vaccination (Cominarty, Spikevax, Vaxzevria with 2-dose scheme and Janssen COVID-19 with a single dose scheme) and mRNA vaccines (Cominarty and Spikevax) are used for the boost(19).

Vaccination exposure, obtained through the electronic nationwide register VACINAS, was classified as: (i) unvaccinated (no register of vaccine administration); (ii) partial primary vaccination (SARS-CoV-2 infection diagnosis less than 14 days after completing the primary vaccination scheme according to the product used); (iii) complete primary vaccination (SARS-CoV-2 infection diagnosis 14 or more days following the complete vaccination scheme according to the product characteristics: 14 days or more days after the second dose of mRNA or Vaxzevria vaccines uptake and 14 days after the single dose of the Janssen COVID-19 vaccine uptake); (iv) partial boost (SARS-CoV-2 infection diagnosis less than 14 days after booster dose uptake); (v) boost complete (SARS-CoV-2 infection diagnosis 14 or more days following booster dose uptake).

To account for time since vaccination uptake, we additionally considered three categories within the complete primary vaccination: (I) complete primary vaccination less than 113 days (16 weeks); (ii) complete primary vaccination 113-168 days (17-24 weeks) and (iii) complete primary vaccination more than 168 days (25 or more weeks).

To avoid small sample size bias, we will not present estimates for vaccine exposure categories with sample size n<20.

### Ethical Statement

This study complied with legal and ethical requirements for research on genomic epidemiology of novel coronavirus (SARS-COV-2).

### Statistical Analysis

Characteristics of participants infected with Omicron and Delta VOC were compared using the chi-square test. Logistic regression adjusted for age group, sex, region of residence, week of diagnosis and laboratory of origin was used to estimate confounding-adjusted odds of complete/boosted vaccination in Omicron cases compared to Delta SARS-CoV-2 cases. An odds ratio (OR) equal to one (OR=1) indicates no difference in odds of vaccination between Omicron and Delta cases and, thus, a proxy of no difference between vaccine effectiveness against Omicron versus Delta VOC. An odds ratio higher than one (OR>1) indicates a higher odds of vaccination among Omicron cases, as such, lower vaccine effectiveness against Omicron compared to Delta VOC. Otherwise an odds ratio lower than one (OR<1) indicates a lower odds of vaccination among Omicron cases, and a higher vaccine effectiveness against Omicron in comparison with Delta VOC.

We also provide estimates of vaccine effectiveness against the Omicron for complete primary vaccination scheme and for the booster dose by combining previously published vaccine effectiveness estimates against Delta and OR estimated in this study using the following formula:

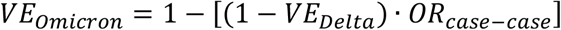

where *VE*_*Omicron*_ represents COVID-19 vaccine effectiveness against Omicron VOC, *VE*_*Delta*_ COVID-19 vaccine effectiveness estimates against Delta VOC and *OR*_*case*−*case*_ the ratio of vaccination odds between Omicron cases versus Delta cases obtain through a case-case design.

To account for uncertainty from the confidence intervals around points estimates we used Monte Carlo simulations, considering that the logarithm of *OR*_*case*−*case*_and logarithm of (1 − *VE*_*Delta*_) are normally distributed. More detailed description of Monte Carlo simulations is provided in supplementary material.

In a secondary analysis, mean and standard deviation (SD) of mean paired Ct values (average of N and ORF) for Omicron and Delta VOC cases were stratified according to vaccination status. Differences between mean Ct values by vaccination status and VOC were evaluated by fitting a linear regression model with Ct values as outcome. Model was adjusted for sex, age group, region, week of case diagnosis, laboratory of origin, vaccination status and VOC type. An interaction term between vaccination status and VOC type was included in the regression model to measure if vaccination status effect on Ct values is differential between Omicron and Delta VOC infection.

### Sensitivity analysis

To assess the bias of misclassification error associated with the SGTF method, we included only cases identified exclusively through WGS. To address if having been infected and vaccinated was associated with lower infectiousness in any of the studied VOCs, we also restricted analysis to the samples with Ct values below 25 (Ct<25) as one may expect that higher Ct values are likely less associated with infectiousness and might frequently yield non-reproducible values.

To account for the effect of previous infection, we performed an additional analysis including cases with previous infection. For this we considered the following levels of exposure, combining information on vaccination status and previous infection: (I) unvaccinated without previous infection; (ii) unvaccinated with previous infection; (iii) partially or completely vaccinated with previous infection; (iv) completely vaccinated without previous infection; (vi) booster vaccination without previous infection. Previous infection was defined as having laboratory confirmation of SARS-CoV-2 at least 90 days prior to current diagnosis by RT-PCR or Rapid antigen test.

## Results

Of a total of 15,001 sample collected during the study period for population aged 12 or more years old, 13,134 were included in the main analysis (Figure1). Of those 4898 (37.3%) were classified as Omicron. Distribution of Omicron cases differed from the distribution of Delta cases by all considered covariates. (Table 1)

**Table 1.** Distribution of SARS-CoV-2 Omicron and Delta cases by method of classification, sex, age, region, week of diagnosis and COVID-19 vaccination status.

|  | Omicron |  | Delta |  | p-value* |
| --- | --- | --- | --- | --- | --- |
|  | n | % | n | % |  |
| **Method of VOC classification** |  |  |  |  |  |
| S-gene | 4732 | 96.6 | 7705 | 93.5 |  |
| WGS | 138 | 2.8 | 452 | 5.5 |  |
| WGS+S-gene | 28 | 0.6 | 88 | 1.1 |  |
| **Sex** |  |  |  |  | 0.009 |
| Female | 2535 | 51.76 | 4074 | 49.41 |  |
| Male | 2363 | 48.24 | 4171 | 50.59 |  |
| **Age group** |  |  |  |  | <0.001 |
| 12-19 | 671 | 13.7 | 648 | 7.9 |  |
| 20-29 | 1508 | 30.8 | 1625 | 19.7 |  |
| 30-39 | 967 | 19.7 | 1779 | 21.6 |  |
| 40-49 | 961 | 19.6 | 1913 | 23.2 |  |
| 50-64 | 745 | 15.2 | 1825 | 22.1 |  |
| 65+ | 46 | 0.9 | 455 | 5.5 |  |
| **Region** |  |  |  |  | <0.001 |
| Alentejo | 258 | 5.3 | 431 | 5.2 |  |
| Algarve | 50 | 1.0 | 153 | 1.9 |  |
| Centro | 414 | 8.5 | 975 | 11.8 |  |
| Norte | 2592 | 52.9 | 5307 | 64.4 |  |
| Lisboa | 1584 | 32.3 | 1379 | 16.7 |  |
| **Week** |  |  |  |  | <0.001 |
| 49 | 184 | 3.8 | 3428 | 41.6 |  |
| 50 | 1032 | 21.1 | 2960 | 35.9 |  |
| 51 | 3682 | 75.2 | 1857 | 22.5 |  |
| **Vaccination Status** |  |  |  |  | <0.001 |
| Unvaccinated | 315 | 6.4 | 888 | 10.8 |  |
| Vaxzevria | 274 | 5.6 | 717 | 8.7 |  |
| Janssen COVID-19 | 876 | 17.9 | 1524 | 18.5 |  |
| Spikevax | 639 | 13.1 | 696 | 8.4 |  |
| Comirnaty | 2794 | 57.0 | 4420 | 53.6 |  |
| **Time since complete vaccination n=11760** |  |  |  |  | <0.001 |
| less than 113 days (16 weeks) | 1002 | 22.2 | 1141 | 15.8 |  |
| 113 to 168 days (16 to 24 weeks) | 2640 | 58.5 | 4313 | 59.5 |  |
| 169 or more days (25 or more weeks) until booster uptake | 873 | 19.3 | 1791 | 24.7 |  |

\*P-value of chi-square test

### Main results

We observed higher odds of complete vaccination in Omicron cases versus Delta (OR=2.1), indicating reduced effectiveness of complete vaccination schemes with mRNA or viral vector vaccines in preventing SARS-CoV-2 infection with the novel Omicron (BA.1) VOC in Portuguese population aged 12 or more years old (Table2).

**Table 2.** Crude and confounder-adjusted odds ratios of vaccine infection breakthrough in Omicron (BA.1) cases compared to Delta (B.1.617.2) SARS-CoV-2 cases, Portugal, weeks 49-51 2021.

| Vaccination status | Omicron<br>n (%) | Delta<br>n (%) | Crude Odds<br>Ratio, (CI95%) | Confounding-adjusted*<br>Odds Ratio,<br>(CI95%) |
| --- | --- | --- | --- | --- |
| **Population aged 12 or more**<br>**years old, n= 13143** | 4898(37.3) | 8245(62.7) |  |  |
| Unvaccinated | 315 (6.4) | 888 (10.8) | ref | ref |
| Partial primary vaccination | 68 (1.4) | 112 (1.4) | 1.7(1.2 to 2.4) | 1.5(0.97 to 2.2) |
| Complete primary vaccination | 4515 (92.2) | 7245 (87.8) | 1.8 (1.5 to 2.0) | 2.1(1.8 to 2.4) |
| Complete primary vaccination<br><113 days | 1002 (22.2) | 1141 (15.5) | 2.5(2.1 to 2.9) | 2.3(1.9 to 2.8) |
| Complete primary vaccination<br>113-168 days | 2640 (58.5) | 4313 (59.5) | 1.7 (1.5 to 2.0) | 2.0 (1.7 to 2.4) |
| Complete primary vaccination<br>169+ days | 873 (19.3) | 1791 (24.7) | 1.4 (1.2 to 1.6) | 1.9(1.6 to 2.3) |
| **Cases with Ct<25(n=11235)** |  |  |  |  |
| Unvaccinated | 261(6.1) | 697(10.1) | ref | ref |
| Partial primary vaccination | 57 (1.3) | 83(1.2) | 1.8(1.3 to 2.6) | 1.7(1.1 to 2.7) |
| Complete primary vaccination | 3979(92.6) | 6158(88.8) | 1.7(1.5 to 2.0) | 2.1(1.8 to 2.6) |
| **Cases classified by WGS (n=706)** |  |  |  |  |
| Unvaccinated | 20 (12.1) | 110 (20.4) | ref | ref |
| Partial primary vaccination | 5 (3.0) | 7 (1.3) | NE | NE |
| Complete primary vaccination | 141 (84.9) | 423 (78.3) | 1.8 (1.1 to 3.1) | 2.7(1.4 to 5.0) |
| **Accounting for previous infection, n=13647** |  |  |  |  |
| Unvaccinated without previous infection | 315 (6.0) | 888 (10.6) | ref | ref |
| Unvaccinated with previous infection | 327(6.2) | 108 (1.3) | 8.5(6.6 to 11.0) | 9.0 (6.6 to 12.3) |
| Partial without previous infection | 68(1.3) | 112 (1.3) | 1.7(1.2 to 2.4) | 1.5 (0.97 to 2.2) |
| Partial/complete with previous infection | 44(0.84) | 25 (0.3) | 5.0(3.0 to 8.2) | 5.1(2.6 to 9.9) |
| Complete without previous infection | 4515(85.7) | 7245 (88.6) | 1.8(1.5 to 2.0) | 2.1 (1.8 to 2.4) |

\*Model adjusted for sex, age group, region, week of diagnosis and laboratory of origin; NE: not estimated due to small sample size

Considering time since complete scheme vaccine uptake, we found statistically significant higher odds of vaccination in Omicron cases for all time intervals since complete vaccination, indicative of VE reduction regardless of time since complete vaccination.

Our results showed higher odds of reinfection with Omicron, regardless vaccination status (Table2) Sensitivity analysis restricted to cases classified only by WGS for the population aged 12 years or older, corroborated the observed differences in odds of complete vaccination between the Omicron and the Delta cases, leading to slightly higher point estimate of OR (OR=2.7). Restriction to samples with Ct<25, also resulted in no relevant change in vaccine breakthrough OR estimates for the population aged 12 years or older.

Our analysis regarding the comparative booster dose vaccine effectiveness against Omicron versus Delta cases was restricted to 3737 cases collected for the population aged 50 or more years old without previous SARS-CoV-2 infection (Table3). Cases characteristics by vaccination status are shown in supplementary material.

**Table 3.**
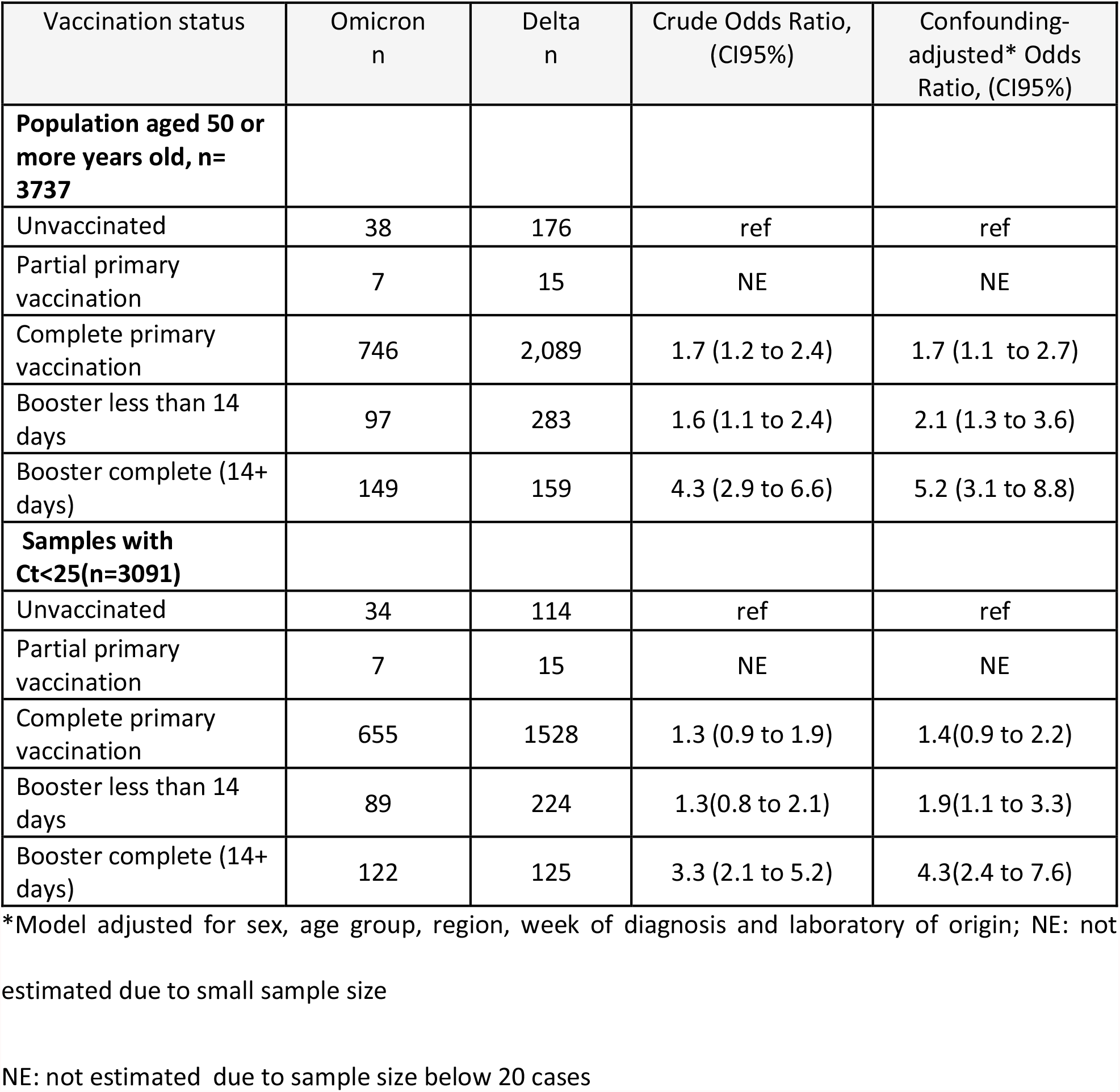
Crude and confounder-adjusted odds ratios of booster dose vaccine infection breakthrough in Omicron (BA.1) cases compared to Delta (B.1.627.2) SARS-CoV-2 cases, Portugal, individuals aged 50 or more years of age, weeks 49-51 2021.

| Vaccination status | Omicron<br>n | Delta<br>n | Crude Odds Ratio,<br>(CI95%) | Confounding-adjusted* Odds<br>Ratio, (CI95%) |
| --- | --- | --- | --- | --- |
| **Population aged 50 or more years old, n= 3737** |  |  |  |  |
| Unvaccinated | 38 | 176 | ref | ref |
| Partial primary vaccination | 7 | 15 | NE | NE |
| Complete primary vaccination | 746 | 2,089 | 1.7 (1.2 to 2.4) | 1.7 (1.1 to 2.7) |
| Booster less than 14 days | 97 | 283 | 1.6 (1.1 to 2.4) | 2.1 (1.3 to 3.6) |
| Booster complete (14+ days) | 149 | 159 | 4.3 (2.9 to 6.6) | 5.2 (3.1 to 8.8) |
| **Samples with Ct<25(n=3091)** |  |  |  |  |
| Unvaccinated | 34 | 114 | ref | ref |
| Partial primary vaccination | 7 | 15 | NE | NE |
| Complete primary vaccination | 655 | 1528 | 1.3 (0.9 to 1.9) | 1.4(0.9 to 2.2) |
| Booster less than 14 days | 89 | 224 | 1.3(0.8 to 2.1) | 1.9(1.1 to 3.3) |
| Booster complete (14+ days) | 122 | 125 | 3.3 (2.1 to 5.2) | 4.3(2.4 to 7.6) |

\*Model adjusted for sex, age group, region, week of diagnosis and laboratory of origin; NE: not estimated due to small sample size
NE: not estimated due to sample size below 20 cases

We observed higher odds of booster vaccination in Omicron cases OR= 5.2 (CI 95%: 3.1 to 8.8) compared to Delta (Table3). The relative reduction of booster vaccine effectiveness was more pronounced compared to the one observed for the complete vaccination in this age group OR=1. 7 (CI 95:1.1 to 2.6).

In sensitivity analysis, restricted to cases with Ct<25 leads to a reduction in OR estimates for both complete primary vaccination (OR=1.4, CI 95: 0.9 to 2.2) scheme and booster vaccination (OR=4.3; Ci 95: 2.4 to 7.6).

Due to small sample size of cases classified by WGS and absence of cases with previous infection among those with booster dose, it was not possible to perform any other sensitivity analysis for population aged 50 or more years old.

### Vaccine effectiveness against Omicron (BA.1) VOC

Using previously published data on VE against symptomatic Delta infection for the primary complete vaccination(20), we estimated VE against infection with the Omicron of 28.1% (CI 95: 12.2 to 40.9%) for the complete primary vaccination scheme and of 68.8% (46.4 to 81.7%) for the booster.

For vaccinated with the Vaxzevria, we obtained VE estimates of -7.1% (CI 95: -31.0 to 11.9) and 64.2% (IC 95: 36.8 to 79.4) for 2-dose primary scheme and for subsequent booster with mRNA vaccine, respectively.

### Ct-values analysis

In our secondary analysis for population aged 12 or more years old for Omicron cases pared mean Ct-values (average of N and ORF) ranged between 18.3 and 18.6 between different vaccination status exposures, while for Delta cases mean Ct-values ranged between 17.8 and 19.5. For overall sample, we observed no statistically significant differences in mean Ct values by VOC (Mean difference (MD): -0.01 (−0.2 to 0.2) (Table 5, Figure S1). Differences in mean Ct-values between Omicron and Delta cases were also not significant for all vaccination exposure levels (unvaccinated, primary partial vaccination, complete primary vaccination). Considering time since complete primary vaccination, we observed differences between Delta and Omicron cases only for cases with less than 113 days since complete primary vaccination (MD=-0.7; CI 95%: -1.13; -0.28), indicating slightly lower Ct (higher viral loads) in Omicron cases.

**Table 4.** Estimates of complete primary scheme COVID-19 Cominarty and Vaxzevria vaccine effectiveness and Comirnaty booster dose vaccine effectiveness against Omicron VOC, based on the combination of VE estimates against Delta VOC previously published(20) and vaccine breakthrough odds ratio infection in Omicron versus Delta VOC cases.

|  | Vaccine effectiveness against Delta VOC (20)* | Odds ratio of vaccine breakthrough infection in Omicron cases versus Delta Cases | Vaccine effectiveness against Omicron VOC |
| --- | --- | --- | --- |
| Comirnaty vaccine |  |  |  |
| Complete vaccine scheme | 62.5% (61.0 to 63.9%) | 2.1(1.8 to 2.4) | 28.1% (12.2 to 40.9%) |
| Booster dose | 94.0% (93.4 to 94.6%) | 5.2 (3.1 to 8.8) | 68.8% (46.4 to 81.7%) |
| Vaxzevria |  |  |  |
| Complete vaccine scheme | 44.1% (41.9 to 46.1%) | 2.1(1.8 to 2.4) | -7.1% (-31.0 to 11.9%) |
| Booster dose of mRNA | 93.1% (91.7 to 94.3%) | 5.2 (3.1 to 8.8) | 64.2% (36.8 to 79.4%) |

**Table 5.**
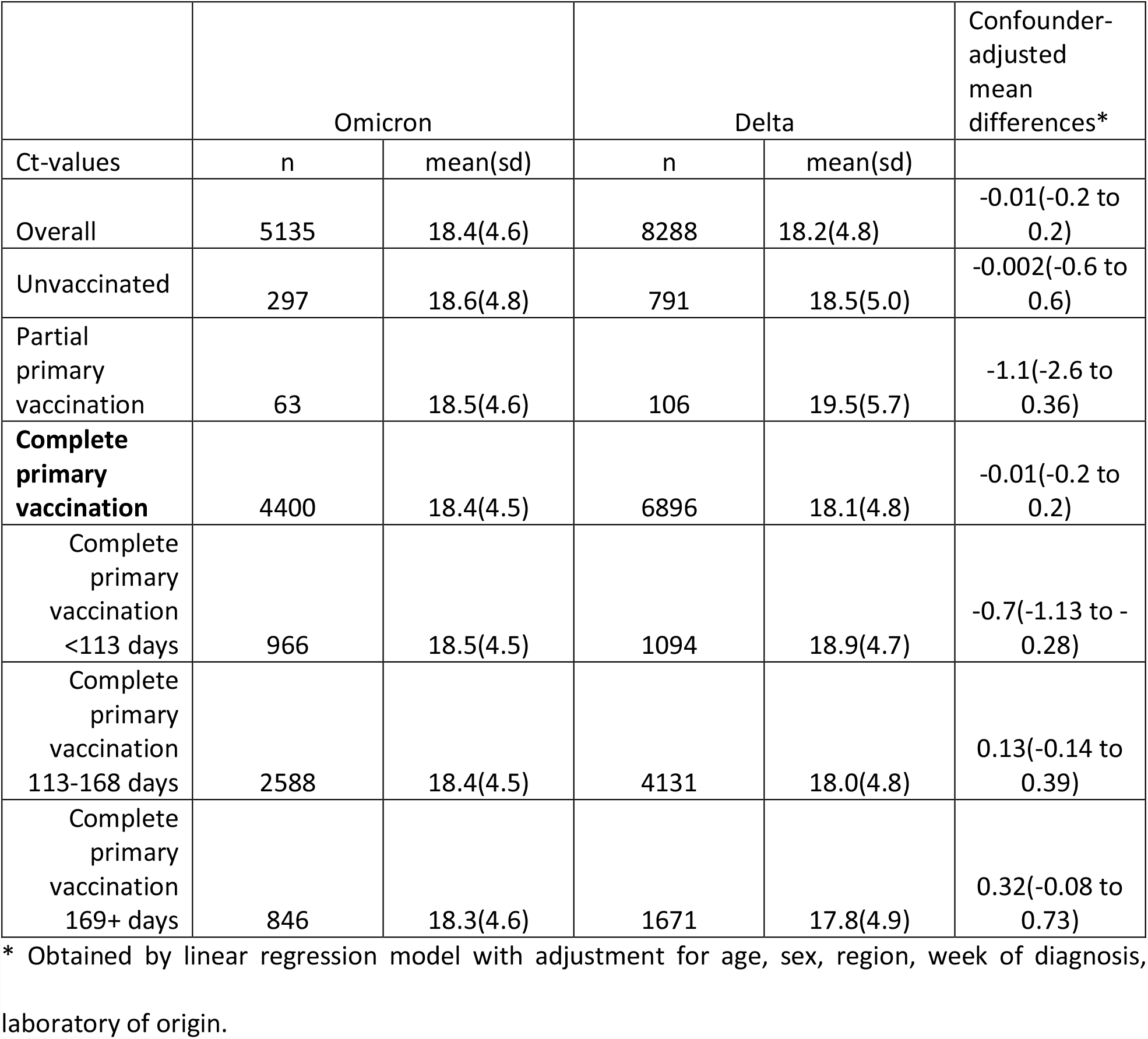
Mean and standard deviation Ct values (N and ORF gene) stratified by vaccination status and VOC and cofounder-adjusted mean differences estimates.

For cases with 50 or more years of age, no statistically significant differences in mean Ct values by VOC or vaccination status were observed, distribution of Ct is shown in supplementary material (Figure S2).

## Discussion

For the Portuguese population aged 12 or more years old, our results show higher odds of complete primary vaccination in cases infected by Omicron (BA.1) VOC compared to Delta (B.1.617.2 and sub-lineages) VOC cases (OR=2.1). This pattern was observed for all time intervals since completion of primary vaccination scheme according to the product characteristics (OR=2.3 for complete vaccination less than 113 days (16 weeks), OR=2.0 for 113-168 days since complete vaccination and OR=1.9 for more than 169 days (25 weeks) since complete vaccination). This finding on consistent difference between Omicron and Delta regardless time since vaccination was similar to primary VE reduction against Omicron VOC observed in UK study(10). Translation of case-case OR estimates to vaccine effectiveness estimates against Omicron VOC infection resulted in substantially lower and even null COVID-19 vaccine effect for complete primary vaccination against Omicron (VE ranged between -7.1% and 28.1%). These results are indicative of low or null protection of complete primary vaccination against SARS-CoV-2 infection also observed in other countries(10–12)

For the booster dose, available at the time of the study for population aged 50 or more years old, we estimated an OR of 5.2 (CI95: 3.1 to 8.8) that also indicated a marked reduction of protection against infection (regardless presence of symptoms) with Omicron compared to Delta. However, since booster has been shown to be highly effective against Delta VOC infection (VE: 94.0 (93.4 to 94.6)), the translation of case-case OR estimates in vaccine effectiveness estimates against Omicron VOC infection, led to estimates ranging from 64.2 to 68.8%. These results were consistent with previous findings from UK study that reported moderate vaccine effectiveness of the booster dose against symptomatic infections with Omicron up to 10 weeks after the booster uptake(10). However, the protection induced by booster dose may be of short term, since considerable VE waning, in particular against SARS-CoV-2 infection, has been previous reported in literature for primary vaccination. So further studies are required to monitor booster VE against Omicron infection along time.

The distributions of Ct-values in our study were similar between VOC and vaccination status. We observed no statistically significant confounding-adjusted mean differences either by vaccination exposure or VOC. Few studies have previously explored the differences in Ct-values between Omicron and Delta VOC. Existing evidence on viral loads differences between Omicron and Delta were not consistent, with studies from Denmark and Switzerland showing similar Ct-value distributions between Delta and Omicron cases(21,22), while in France significantly higher Ct values were observed for cases infected with Omicron(23). No significant differences in the Ct values between people infected with Omicron and people infected with Delta is thus not surprising and it has been corroborated by other studies(21,22). In fact, it is believed that the molecular basis underlying Omicron’s higher transmissibility likely relies on its mutational pattern involved in alternate entry mechanisms, increased host-cell binding and immune evasion, rather on higher viral loads spread through aerosolization(24,25).

Our study has several limitations. First, we were unable to differentiate between symptomatic and asymptomatic infections. Second, our study was based on RT-PCR tests, so rapid antigen test widely used in Portugal for SARS-CoV-2 diagnosis(26) were not covered by our data. Although study included cases samples from National SARS-CoV-2 Genomic Surveillance Network and three major clinical pathology laboratories, we cannot exclude that cases included in our study might not be representative of the overall infections detected in Portugal during week 49-51 2021. Hence, we compared age group, sex and region of study sample and the overall cases notified with infection in the same period. We found no significant differences in the sex and age group distribution between the universe of identified cases through rapid or RT-PCR testing and notified to SINAVE and the study sample. However, the north region tends to be overrepresented in our study sample. Third, incentives for testing had changed during the study period, negative tests regardless vaccination status became required to go to restaurants, hotels and to participate cultural events(27). This change may affect study results. However, selection of cases to the study sample was independent from the VOC type and from vaccination status. Fourth, this study did not collected data on comorbidities, considered relevant confounding variables in vaccines effectiveness research. So, we were not able to adjust for comorbidities in our models, neither estimate proportion of those with comorbidities by vaccination status. This can affect study results since individuals with comorbidities, such as immunosuppressed patients were prioritized for additional vaccination dose in Portugal(19). Finally, to translate case-case result to VE estimates we used previously published estimates of VE against symptomatic infection with Delta, while our study sample included notified infections regardless their symptoms.

Among study strengths, we should mention its large sample size due to use of surveillance data, well-established and high positive predictive value in SGTF method to classy infections as Delta or Omicron variants, and the robustness of results when we changed the sampling strategy in our sensitivity analyses (WSG-only or restriction to Ct<25).

Although direct comparisons to other studies are challenging due to differences in methodology, outcome and exposure definitions, vaccination calendar, eligibility for booster, vaccine brand-specific policies, testing patterns and other non-pharmacological interventions in place, our findings corroborate the general trend. More specifically our results support a marked reduction of primary and booster vaccination schemes effectiveness in preventing Omicron infections compared to Delta observed in other countries(10,12). Our findings suggest that complete primary vaccination may not be protective against SARS-CoV-2 infection in regions where Omicron variant is dominant. Even with lower booster vaccine effectiveness against Omicron, massive roll out and acceleration of booster vaccination campaign can contribute to reduction of number of susceptible to SARS-CoV-2 infection and its incidence in the population.

## Data Availability

data can be provided by the data owners upon reasonable request

## Supplementary data

**Table S1.** Distribution of SARS-CoV-2 Omicron (BA.1) and Delta (B1.617.2) cases by method of classification, sex, age, region, week of diagnosis and COVID-19 vaccination status for 50 or more years old

|  | Omicron |  | Delta |  |  |
| --- | --- | --- | --- | --- | --- |
|  | n | % | n | % | p-value |
| Sample type |  |  |  |  | <0.001 |
| S-gene | 998 | 96.9 | 2431 | 89.8 |  |
| WGS | 28 | 2.7 | 243 | 9.0 |  |
| WGS+S-gene | 4 | 0.4 | 33 | 1.2 |  |
| Sex |  |  |  |  | 0.293 |
| Female | 522 | 50.7 | 1424 | 52.6 |  |
| Male | 508 | 49.3 | 1283 | 47.4 |  |
| Age group |  |  |  |  | <0.001 |
| 50-64 | 837 | 81.3 | 1945 | 71.9 |  |
| 65+ | 193 | 18.7 | 762 | 28.2 |  |
| Region |  |  |  |  | <0.001 |
| Alentejo | 73 | 7.1 | 162 | 6.0 |  |
| Algarve | 11 | 1.1 | 65 | 2.4 |  |
| Centro | 90 | 8.7 | 367 | 13.6 |  |
| Norte | 519 | 50.4 | 1756 | 64.9 |  |
| AM Lisboa | 337 | 32.7 | 357 | 13.2 |  |
| Week |  |  |  |  | <0.001 |
| 49 | 45 | 4.4 | 1178 | 43.5 |  |
| 50 | 190 | 18.5 | 949 | 35.1 |  |
| 51 | 795 | 77.2 | 580 | 21.4 |  |
| Vaccination Status |  |  |  |  | <0.001 |
| Unvaccinated | 38 | 3.7 | 176 | 6.5 |  |
| Covid19AstraZeneca | 197 | 19.1 | 581 | 21.5 |  |
| Covid19Janssen | 90 | 8.7 | 282 | 10.4 |  |
| Covid19Moderna | 117 | 11.4 | 236 | 8.7 |  |
| Covid19Pfizer | 588 | 57.1 | 1432 | 52.9 |  |

**Figure S1.**
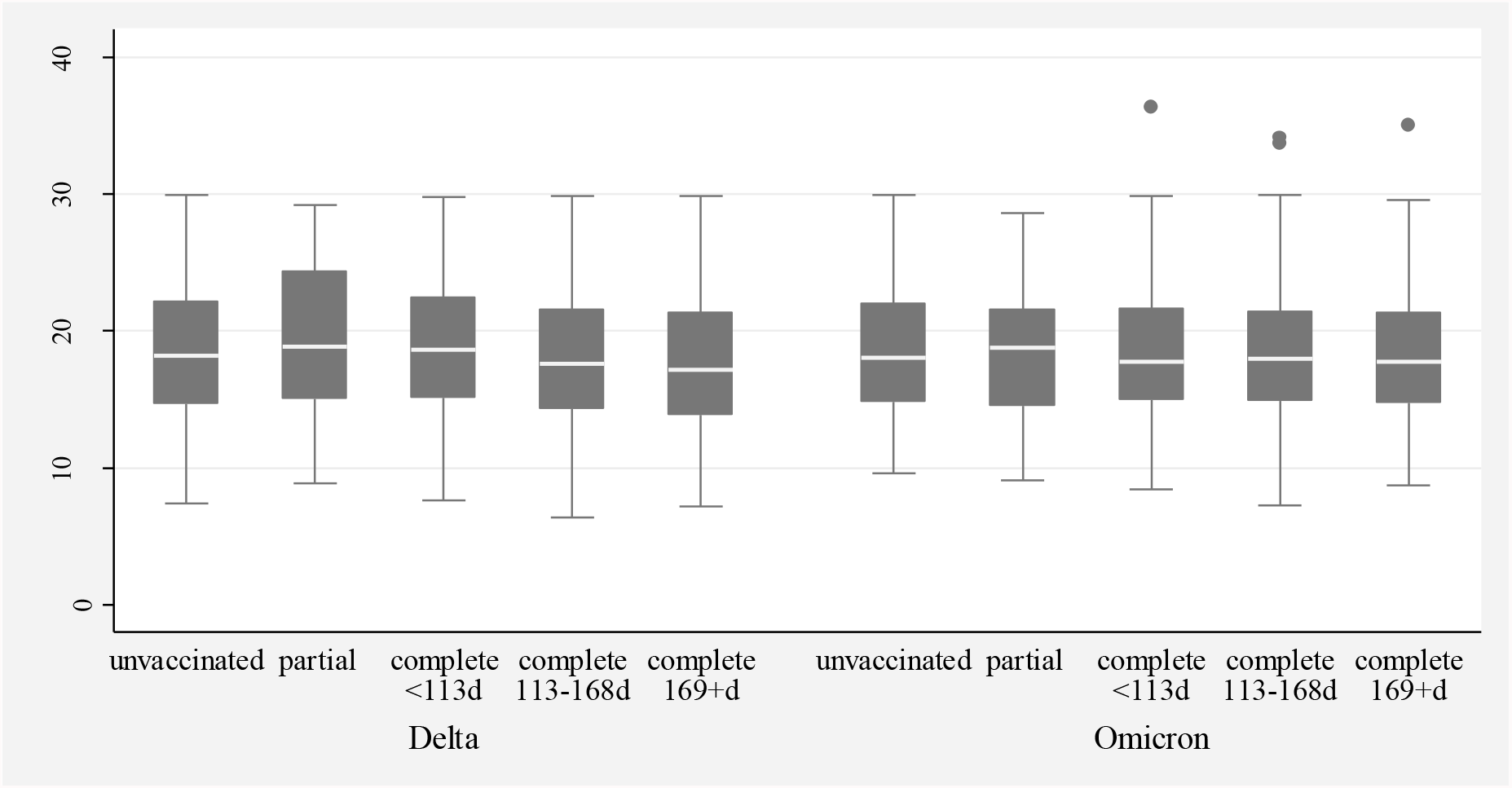
Distribution of Ct values by VOC and vaccination status, population aged 12 or more

**Figure S2.**
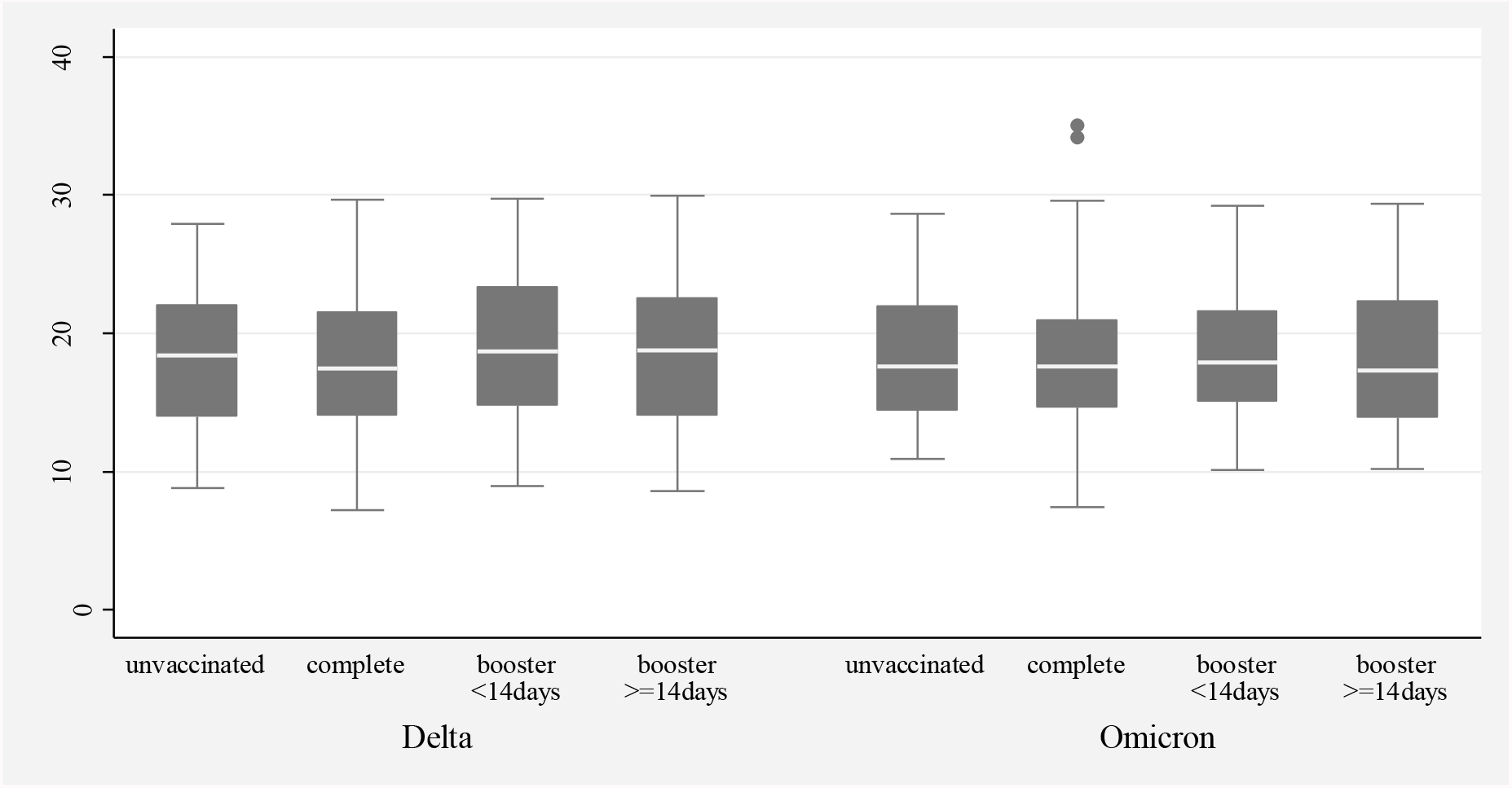
Distribution of Ct values by VOC and vaccination status, population aged 50 or more

## Construction of empirical distributions and estimation of confidence intervals for COVID-19 vaccine effectiveness (VE) against Omicron using data on VE against Delta and odds ratio from case-case study design

We computed estimates of vaccine effectiveness against the Omicron for complete primary vaccination scheme and for the booster dose by combining previously published vaccine effectiveness estimates against Delta and OR estimated in this study using the following formula:

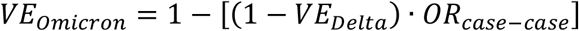

where *VE*_*Omicron*_ represents COVID-19 vaccine effectiveness against Omicron VOC, *VE*_*Delta*_ COVID-19 vaccine effectiveness estimates against Delta VOC and *OR*_*case*−*case*_ the ratio of vaccination odds between Omicron cases versus Delta cases obtain through a case-case design. To estimate 95% confidence intervals for COVID-19 VE against Omicron we used Monte Carlo simulations. First, we constructed empirical distributions for all input parameters, namely of vaccine effectiveness against Delta (*VE*_*Delta*_) and *OR*_*case*−*case*_ from case-case design. To construct empirical distribution of *VE*_*Delta*_ we assumed a normal distribution for log(1-*VE*_*Delta*_), since COVID-19 *VE*_*Delta*_ estimates were obtained in test-negative design study as *VE*_*Delta*_=1-Odds ratio(*OR*_*Delta*_), estimated by logistic regression model, as *OR*_*Delta*_=exp(*β*_*ve*_*delta*_), where *β*_*ve*_*delta*_ represents coefficient from logistic regression model for vaccination exposure. We transformed *VE*_*Delta*_ into 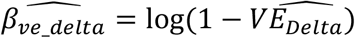 and *VE*_*Delta*_ 95% confidence interval upper 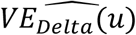 and lower 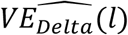 bounds into 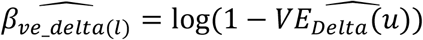 and 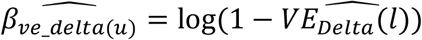, respectively. We computed a standard error 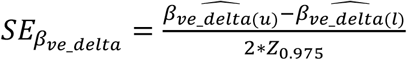, where *Z*_0.975_ represents a quantile of standard Normal distribution, and generated pseudo-random numbers from Normal distribution:

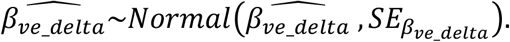

Simulated values of 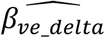 were transformed back to original scale using following formula: 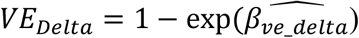.

To construct empirical distribution of *OR*_*case*−*case*_we generated pseudo-random numbers from Normal distribution 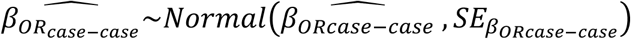 where 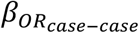 represents coefficient from logistic regression model and 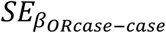 its standard error

To return to original scale, we applied an inverse transformation:

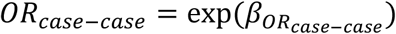

We draw 10 000 samples of *OR*_*Delta*_ and *VE*_*Delta*_ used them to construct empirical distributions of *VE*_*Omicron*_

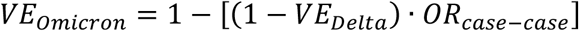

The 2.5 and 97.5 percentiles of these empirical distributions were used as lower and upper limits of the 95% confidence intervals for *VE*_*Omicron*_

## References

1. Viana R, Moyo S, Amoako DG, Tegally H, Scheepers C, Althaus CL, et al. Rapid epidemic expansion of the SARS-CoV-2 Omicron variant in southern Africa. medRxiv [Internet]. 2021 Dec 21 [cited 2022 Jan 31];18:2021.12.19.21268028. Available from: https://www.medrxiv.org/content/10.1101/2021.12.19.21268028v1

2. World Health Organization. Classification of Omicron (B.1.1.529): SARS-CoV-2 Variant of Concern [Internet]. Vol. 337, WHO. 2021 [cited 2022 Jan 31]. Available from: https://www.who.int/news/item/26-11-2021-classification-of-omicron-(b.1.1.529)-sars-cov-2-variant-of-concern

3. Pulliam JRC, Schalkwyk C van, Govender N, Gottberg A von, Cohen C, Groome MJ, et al. Increased risk of SARS-CoV-2 reinfection associated with emergence of the Omicron variant in South Africa. medRxiv [Internet]. 2021 Dec 2 [cited 2022 Jan 31];2021.11.11.21266068. Available from: https://www.medrxiv.org/content/10.1101/2021.11.11.21266068v2

4. Karim SSA, Karim QA. Omicron SARS-CoV-2 variant: a new chapter in the COVID-19 pandemic. Lancet (London, England) [Internet]. 2021 Dec [cited 2022 Jan 31];398(10317):2126–8. Available from: https://pubmed.ncbi.nlm.nih.gov/34871545/

5. European Centre for Disease Prevention and Control (ECDC). Methods for the detection and characterisation of SARS-CoV-2 variants - first update 20 Dec 2021. 2021;

6. Espenhain L, Funk T, Overvad M, Edslev SM, Fonager J, Ingham AC, et al. Epidemiological characterisation of the first 785 SARS-CoV-2 Omicron variant cases in Denmark, December 2021. Eurosurveillance [Internet]. 2021 Dec 16 [cited 2022 Jan 31];26(50):2101146. Available from: https://www.eurosurveillance.org/content/10.2807/1560-7917.ES.2021.26.50.2101146

7. European Centre for Disease Prevention and Control (ECDC). Threat Assessment Brief: Implications of the emergence and spread of the SARS-CoV-2 B.1.1. 529 variant of concern (Omicron) for the EU/EEA [Internet]. 2021 [cited 2022 Jan 31]. Available from: https://www.ecdc.europa.eu/en/publications-data/covid-19-threat-assessment-spread-omicron-first-update

8. Cele S, Jackson L, Khoury DS, Khan K, Moyo-Gwete T, Tegally H, et al. SARS-CoV-2 Omicron has extensive but incomplete escape of Pfizer BNT162b2 elicited neutralization and requires ACE2 for infection. medRxiv Prepr Serv Heal Sci [Internet]. 2021 Dec 17 [cited 2022 Jan 31]; Available from: https://pubmed.ncbi.nlm.nih.gov/34909788/

9. Gruell H, Vanshylla K, Tober-Lau P, Hillus D, Schommers P, Lehmann C, et al. mRNA booster immunization elicits potent neutralizing serum activity against the SARS-CoV-2 Omicron variant. Nat Med 2022 [Internet]. 2022 Jan 19 [cited 2022 Jan 31];1–4. Available from: https://www.nature.com/articles/s41591-021-01676-0

10. Andrews N, Stowe J, Kirsebom F, Toffa S, Rickeard T, Gallagher E, et al. Effectiveness of COVID-19 vaccines against the Omicron (B.1.1.529) variant of concern. medRxiv [Internet]. 2021 Dec 14 [cited 2022 Jan 31];2021.12.14.21267615. Available from: https://www.medrxiv.org/content/10.1101/2021.12.14.21267615v1

11. Sheikh A, Kerr S, Mcmenamin J, Robertson C. Severity of Omicron variant of concern and vaccine effectiveness against symptomatic disease: national cohort with nested test negative design study in Scotland.

12. Hansen CH, Schelde AB, Moustsen-Helm IR, Emborg H-D, Krause TG, Mølbak K, et al. Vaccine effectiveness against SARS-CoV-2 infection with the Omicron or Delta variants following a two-dose or booster BNT162b2 or mRNA-1273 vaccination series: A Danish cohort study. medRxiv [Internet]. 2021 Dec 23 [cited 2022 Jan 31];2021.12.20.21267966. Available from: https://www.medrxiv.org/content/10.1101/2021.12.20.21267966v3

13. Kislaya I, Rodrigues EF, Borges V, Gomes JP, Sousa C, Almeida JP, et al. Comparative Effectiveness of Coronavirus Vaccine in Preventing Breakthrough Infections among Vaccinated Persons Infected with Delta and Alpha Variants - Volume 28, Number 2— February 2022 - Emerging Infectious Diseases journal - CDC. Emerg Infect Dis [Internet]. 2022 Feb [cited 2022 Jan 31];28(2):331–7. Available from: https://www.nc.cdc.gov/eid/article/28/2/21-1789_article

14. Pogreba-Brown K, Austhof E, Ellingson K. Methodology minute: An overview of the case–case study design and its applications in infection prevention. Am J Infect Control [Internet]. 2020 [cited 2021 Jun 28];48(3):342–4. Available from: https://reader.elsevier.com/reader/sd/pii/S019665531930759X?token=7101205F95AD08427009BCCBD323BC6EAA3D394AD73CE961ADD0069A88EF5D2AAF1AD48EC129092AD96E36E8127FA30C&originRegion=eu-west-1&originCreation=20210628081747

15. Rabaan AA, Tirupathi R, Sule AA, Aldali J, Mutair A Al, Alhumaid S, et al. Viral dynamics and real-time rt-pcr ct values correlation with disease severity in covid-19. Vol. 11, Diagnostics. 2021. p. 1091.

16. Borges V, Sousa C, Menezes L, Gonçalves AM, Picão M, Almeida JP, et al. Tracking SARS-CoV-2 lineage B.1.1.7 dissemination: insights from nationwide spike gene target failure (SGTF) and spike gene late detection (SGTL) data, Portugal, week 49 2020 to week 3 2021. Euro Surveill [Internet]. 2021 Mar 1 [cited 2022 Jan 31];26(10):1–6. Available from: https://pubmed.ncbi.nlm.nih.gov/33706862/

17. Instituto Nacional de Saúde Dr. Ricardo Jorge. Diversidade genética do novo coronavírus SARS-CoV-2 (COVID-19) em Portugal. 28.12.2021 [Internet]. Lisboa; 2021. Available from: https://insaflu.insa.pt/covid19/relatorios/INSA_SARS_CoV_2_DIVERSIDADE_GENETICA_relatorio_situacao_2021-12-28.pdf

18. Instituto Nacional de Saúde Dr. Ricardo Jorge. Diversidade genética do novo coronavírus SARS-CoV-2 (COVID-19) em Portugal. 11.01.2022. 2022.

19. Direção-Geral da Saúde. Norma 002/2021. Campanha de Vacinação Contra a COVID-19 [Internet]. 2021. Available from: https://covid19.min-saude.pt/wp-content/uploads/2021/05/i027514.pdf

20. Andrews N, Stowe J, Kirsebom F, Gower C, Ramsay M, Bernal JL. Effectiveness of BNT162b2 (Comirnaty, Pfizer-BioNTech) COVID-19 booster vaccine against covid-19 related symptoms in England: test negative case-control study. medRxiv [Internet]. 2021 Nov 15 [cited 2022 Jan 31];2021.11.15.21266341. Available from: https://www.medrxiv.org/content/10.1101/2021.11.15.21266341v1

21. Puhach O, Adea K, Hulo N, Sattonnet P, Genecand C, Iten A, et al. Infectious viral load in unvaccinated and vaccinated patients infected with SARS-CoV-2 WT, Delta and Omicron. medRxiv [Internet]. 2022 Jan 18 [cited 2022 Jan 31];2022.01.10.22269010. Available from: https://www.medrxiv.org/content/10.1101/2022.01.10.22269010v2

22. Lyngse FP, Mortensen LH, Denwood MJ, Christiansen LE, Møller CH, Skov RL, et al. SARS-CoV-2 Omicron VOC Transmission in Danish Households. medRxiv [Internet]. 2021 Dec 27 [cited 2022 Jan 31];2021.12.27.21268278. Available from: https://www.medrxiv.org/content/10.1101/2021.12.27.21268278v1

23. Sofonea MT, Roquebert B, Foulongne V, Verdurme L, Trombert-Paolantoni S, Roussel M, et al. From Delta to Omicron: analysing the SARS-CoV-2 epidemic in France using variant-specific screening tests (September 1 to December 18, 2021). medRxiv [Internet]. 2022 Jan 1 [cited 2022 Jan 31];2021.12.31.21268583. Available from: https://www.medrxiv.org/content/10.1101/2021.12.31.21268583v1

24. Zhang L, Li Q, Liang Z, Li T, Liu S, Cui Q, et al. The significant immune escape of pseudotyped SARS-CoV-2 variant Omicron. Emerg Microbes Infect [Internet]. 2022 Dec 31 [cited 2022 Jan 31];11(1):1. Available from: /pmc/articles/PMC8725892/

25. Peacock TP, Brown JC, Zhou J, Thakur N, Newman J, Kugathasan R, et al. The SARS-CoV-2 variant, Omicron, shows rapid replication in human primary nasal epithelial cultures and efficiently uses the endosomal route of entry. bioRxiv [Internet]. 2022 Jan 3 [cited 2022 Jan 31];2021.12.31.474653. Available from: https://www.biorxiv.org/content/10.1101/2021.12.31.474653v1

26. Direção-Geral de Saúde. Norma 015/2020 COVID-19: Rastreio de Contactos. 2021.

27. Resolução do Conselho de Ministros n.o 181-A/2021 | DRE Medidas aplicáveis no âmbito da pandemia da doença COVID-19. [Internet]. Available from: https://dre.pt/dre/detalhe/resolucao-conselho-ministros/181-a-2021-176492317

